# Exploring self-reported quality of life in developmental language disorder

**DOI:** 10.1101/2024.06.19.24309096

**Authors:** Caroline Larson

## Abstract

**Purpose:** Developmental language disorder (DLD) is a lifelong condition associated with poorer outcomes than neurotypical peers, yet relatively little is known about long-term quality of life in DLD. This preliminary study adopts a neurodiversity-informed approach by exploring *self-reported* quality of life in an adolescent and young adult DLD sample, as well as linguistic and risk factors contributing to quality of life.

**Method:** Participants were five individuals with DLD aged 12-20 years (M = 15.60; SD = 3.05). I administered two self-report quality of life scales, a language assessment, an experimental morphosyntax task, and measures of risk factors. Data were analyzed descriptively.

**Results:** Participants generally reported positive views about their quality of life, though accessing accommodations and health services emerged as barriers. Relatively better grammaticality judgement performance appeared to be linked with poorer ratings of happiness and the ability to ‘be yourself.’ Nonverbal ability represented a potential risk factor, though there may be a stronger cumulative role for risk factors.

**Conclusions:** DLD participants reported relatively good quality of life. Exploratory findings suggest barriers to quality of life in some contexts, as well as roles for individual differences in language and risk factors. These descriptive findings should be examined in larger scale studies and may represent areas of consideration when clinicians address functional challenges that impact mental health and wellbeing in individuals with DLD.

---

Developmental language disorder (DLD) is a highly prevalent (Bishop et al., 2017; Norbury et al., 2016) and lifelong neurodevelopmental condition (Botting & Botting, 2020; McGregor, 2020; Nippold & Schwarz, 2002; Tomblin et al., 1992; Whitehouse et al., 2009). It is associated with poorer long-term academic, vocational, and social-emotional outcomes than neurotypical peers (NT; Conti-Ramsden et al., 2013; Conti-Ramsden & Durkin, 2012; Durkin et al., 2012, 2015; Le et al., 2021; Records et al., 1992; Whitehouse et al., 2009; Ziegenfusz et al., 2022). Historically, much of the research on functioning in neurodevelopmental conditions, like DLD, has focused on childhood rather than adolescence and adulthood. Recently, however, there have been calls for research that treats neurodevelopmental conditions as persistent and having important impacts on functioning through adulthood (Antolini & Colizzi, 2023). There have also been recent calls for neurodiversity-informed evidence that characterizes the lived experiences of individuals with neurodevelopmental conditions, particularly evidence obtained from *self-report* in addition to parent/caregiver or teacher report (e.g., Eigsti et al., 2023; Georgiades & Kasari, 2018; Hobson et al., 2024; Orrego et al., 2023). An increased focus on quality of life indicators related to everyday functioning has emerged in recent years (Eadie et al., 2018; Haukedal et al., 2023), as well as first-hand accounts of lived experiences that describe relationships, supports available across development, and other self-reported successes and challenges (e.g., in DLD, Orrego et al., 2023, and in autism, Kim, 2019). Though there are a few studies published in the last 30 years on self-reported quality of life, they are limited to studies from one project that took place in the United Kingdom (Conti-Ramsden et al., 2017) and an early study by Tomblin and colleagues (Records et al., 1992). Examining quality of life indicators derived from first-hand accounts of lived experiences can further reveal how DLD unfolds across the lifespan. The current descriptive data will contribute to a better understanding of how individual differences in the presentation of DLD may affect long-term mental health and wellbeing, both of which are global health concerns (CDC, 2020; Connell et al., 2014), providing guidance for future, larger-scale research.

The current literature using *self-report* quality of life measures in adolescents and adults with DLD appears to include **only three studies**. Records et al. (1992) compared composite measures of quality of life in adolescents and adults with DLD to NT peers and found no significant group differences on measures including general affect (e.g., positive versus negative feelings about life), satisfaction with life (e.g., satisfaction with friendships), and perceived internal/external locus of control (i.e., subjective degree to which control of one’s life is perceived as within the self) after controlling for multiple statistical comparisons. Yet, general affect and locus of control were lower in the DLD group, suggesting lower quality of life in at least some domains. Conti-Ramsden et al. (2013) reported more problems with peer relationships, emotional symptoms, hyperactivity, and conduct problems in their adolescent DLD group compared to NT peers. Each of these issues may contribute to poorer quality of life and are perceived as functional challenges. In a smaller sample from the same project, Durkin et al. (2012) did not find DLD-NT group differences in emotional health (e.g., depression, anxiety) for younger adolescents with DLD, though the DLD group also did not differ from autistic peers who are known to frequently experience emotional health challenges. Thus, the interpretation from the broader Conti-Ramsden et al. (2017) sample suggests that individuals with DLD report at least some emotional health challenges which likely affect their quality of life and may reflect lower ratings of general affect observed in Records et al. (1992). However, Records et al. (1992) is the only study, to my knowledge, that directly examines *self-reported* quality of life outcomes in adolescents and young adults with DLD.

Evidence from *parent-report* suggests that children with DLD experience challenges in quality of life and wellbeing which may worsen across development (Eadie et al., 2018; Le et al., 2021). Using the parent-report Pediatric Quality of Life Inventory, three studies have shown poorer quality of life scores across the domains of physical health, and emotional, social, and school functioning than NT peers (Eadie et al., 2018; Haukedal et al., 2023; Le et al., 2021). These studies also demonstrated that language skills were associated with concurrent (Haukedal et al., 2023; Le et al., 2021) and later (Eadie et al., 2018) quality of life. Toseeb et al. (2023) reported that adolescents with DLD were more likely than NT peers to experience functional challenges in mental health, and that risk factors, including socioeconomic status, family history of psychiatric conditions, and the early language and communication home environment, were important determiners in the degree of functional challenge. McGregor et al. (2023) found that parent-reported challenges in communication, relationships, and academic functioning in children with DLD were associated with risk factors, such as child nonverbal abilities, child health, and caregiver education, but not with individual differences in language skills. This study also suggested relative strengths in daily living skills, play and coping in social contexts, and prosocial qualities (e.g., agency and resilience; see Conti-Ramsden et al., 2013 for converging evidence on prosocial behavior).

Additionally, there may be discrete pathways of influence between the DLD profile and particular outcomes. For instance, in one of the only studies to use parent- *and* self-report measures of outcomes, Durkin et al. (2012) demonstrated an association between language skills and educational achievement in a sample of adolescents who had DLD or autism and co-occurring language impairment. There was no significant association between language skills and anxiety or depression at 14 years of age, and no significant association between language skills and independence or friendships at 14 or 16 years of age when covarying autism symptomology. In a follow-up study from the same group with a larger DLD-only sample of 16-year-olds, language skills were associated with self-reported prosocial behavior, hyperactivity, conduct, and emotional problems, but not peer problems, when covarying gender and nonverbal abilities (Conti-Ramsden et al., 2013). These patterns may reflect relationships between language and quality of life in DLD that are not present in autistic peers with language impairment, though this work did not directly examine self-reported quality of life.

The role of language and other risk factors in quality of life may also vary depending on functional demands associated with given developmental stages. In their first-person, descriptive account of lived experiences of an adult with DLD, Orrego et al. (2023) suggests that functional challenges this individual faced varied depending on changing expectations of her environment. Examples are social barriers related to making friends and getting bullied in elementary school versus educational barriers related to diagnosis disclosure and receiving accommodations in college. These experiences may not be captured in studies that rely on *parent report* or in studies that include only traditional, decontextualized measures of outcomes. Measures that capture lived experiences, like self-reported quality of life indicators, can provide further insight and represent a neurodiversity-informed approach. Orrego et al. (2023) underscores recent recommendations to characterize the clinical course of a condition across the lifespan. Examining quality of life in transitional phases like adolescence through young adulthood is critical because mental health and wellbeing are especially vulnerable during these developmental periods (Antolini & Colizzi, 2023). Taken together, the current state of the literature on self-reported quality of life in adolescents and young adults with DLD is in a nascent stage, and there is **no prior research** that examines the role of individual differences in language or risk factors. This preliminary study contributes novel descriptive evidence on self-reported quality of life in a small adolescent and young adult DLD sample, and linguistic and risk factors that contribute to wellbeing.

## Methods

This study was approved by the University of Connecticut Institutional Review Board and all participants consented to participate in the study. DLD participants were primarily referred to the study by parents, except for participant 5 (P5) who referred themself due to experiencing language-based challenges in college. Participants were five individuals age 12-20 years with DLD (M = 15.60; SD = 3.05) as this age span provides information about different stages of adolescence/adulthood; participants capture the developmental span from early high school to early college. DLD group eligibility was based on current concerns about language skills, a history of specialized intervention related to language or literacy skills, standardized scores <85 on at least one composite scale from the Clinical Evaluation of Language Fundamentals Fifth Edition (CELF-5; Wiig et al., 2013), and matrix reasoning *t*-scores > 70 on the Wechsler Abbreviated Intelligence Scale (WASI; Wechsler, 2011). One DLD participant, P2, had a Core Language score of 87 yet had a scaled score of 5 on the understanding spoken paragraphs subscale and a scaled score of 7 on the word definitions subscale (word definitions is one key component for diagnosis of DLD in adulthood; Fidler et al., 2011). This participant was included due to history of receiving speech-language and special education services for language-based reading difficulty, as well as current parent-reported language concerns (see McGregor et al., 2017 for evidence of adults with DLD scoring within 1 standard deviation of the mean on standardized language assessments and Tomblin et al., 1992 for additional methods identifying DLD in adulthood). All DLD participants were currently receiving or had a history of specialized services for language-based challenges, such as reading or listening comprehension, suggesting the presence of language deficits and previously un-identified DLD in some cases (Adlof & Hogan, 2018; see Wittke & Spaulding, 2018, for evidence related to limitations in the identification of DLD and Mcgregor, 2020, for discussion of individuals with DLD being underserved). See Table 1 for by-participant descriptive characteristics and Supplementary Materials Tables 1 and 2 for additional, group-level descriptive data.

**Table 1.** By-participant descriptive data.

|  | P1 | P2 | P3 | P4 | P5 |
| --- | --- | --- | --- | --- | --- |
| CELF-5 |  |  |  |  |  |
| *Core Language* | 65 | 87 | 83 | NA | 84 |
| *Expressive Language* | 73 | 100 | 83 | 80 | 83 |
| *Receptive Language* | 73 | 92 | 92 | NA | 98 |
| *Language Content Index* | 80 | 96 | 90 | 80 | 100 |
| *Recalling sentences* | 4 | 8 | 7 | 7 | 9 |
| *Word definitions* | 3 | 7 | 8 | 7 | 12 |
| *Understanding spoken paragraphs* | 5 | 5 | 7 | 4 | 7 |
| *Formulated sentences* | 5 | 10 | 7 | 6 | 6 |
| Grammaticality Judgement A` | 0.53 | 0.52 | 0.61 | 0.66 | 0.52 |
| WASI Matrix Reasoning | 41 | NA | 53 | NA | 38 |
| Current education completed (current education placement) | In high school | In high school | In high school | In high school | In college |

*Note.* P = Participant; Core, Expressive, and Receptive Language and Language Content Index = Standard scores; Recalling Sentences, Formulated Sentences, and Understanding Spoken Paragraphs = Scaled scores; Grammaticality Judgement A` = A prime calculated per signal detection theory; WASI = Wechsler Abbreviated Intelligence Scale, 2<sup>nd</sup> Edition, *t*-scores.

### Measures

#### Grammaticality Judgement

This experimental task tests a key area of disproportionate weakness in DLD, morphosyntax. There were 23 grammatical and 23 ungrammatical pseudo-randomized sentences. Ungrammatical sentences contained errors of morphosyntax per General American English, including word order, omission, substitutions, and tense marking. Stimuli were presented auditorily from recordings of a native-English speaker, and participants made a two alternative forced choice judgement of correct (grammatical) or incorrect (ungrammatical). We examined performance according to signal detection theory, using A’ (A prime) to measure participants’ ability to detect grammatical sentences separate from response bias.

#### Quality of Life

Quality of Life was measured using the ASD-Quality of Life Scale (McConachie et al., 2017) involving 9 questions using a 5-point likert scale. Each question was analyzed individually to yield a sensitive measurement of different factors contributing to quality of life. We also measured life satisfaction as another, more general quality of life indicator using the Satisfaction with Life Scale (Diener et al., 1985), which involves 7 questions about life satisfaction using a 7-point likert scale. Higher ratings suggest more life satisfaction; scores were aggregated across the 7 questions to provide a general outcome measure. Measures were participant self-report and administered via Qualtrics surveys. In general, scores above the mean likert value (e.g., ≥3) suggest relatively good quality of life.

#### Risk Factors

Risk factors were based on McGregor et al. (2023) and Toseeb et al. (2023). Participants and parents of participants completed sociodemographic background history questionnaires on gender, race, ethnicity, and socioeconomic status based on the MacArthur Network on Socioeconomic Status and Health (Adler, 2007), administered via Qualtrics surveys. Socioeconomic status was measured by parent reported family income and caregiver highest education. Background history included information on additional relevant risk factors, including parent-reported health history. Nonverbal abilities measured by the WASI also represented a risk factor. See Supplementary Materials Table 2 for sociodemographic characteristics, and Table 1 and Supplementary Materials Tables 3 and 5 for DLD by-participant risk factors.

### Analysis

Descriptive analysis was conducted to explore participant responses to quality of life measures, and the roles of risk factors in quality of life were analyzed by descriptively linking risk factors with quality of life responses for each participant (see Supplementary Materials Table 4 for standards on reporting such methods and additional details on the descriptive approach).

## Results

### Quality of Life

DLD participants reported relatively strong support from others in making decisions and in generally considering themselves a happy person (Table 2). They also indicated relatively good satisfaction with their lives, such as getting important things they want in life and having excellent life conditions. However, there were some indications of lower happiness ratings and barriers to participating in their environments. P2, P3, and P5 reported feeling happy a somewhat low percentage of the time (≤55% of the time). P3, P4, and P5 reported frequently experiencing sensory-based barriers in the environment and barriers to their needs being met in official situations (e.g., interacting with authority figures at school or work, disclosing their diagnosis). When describing barriers to their needs being met in more detail, P3 indicated that they have difficulty accessing school-based accommodations due to teacher resistance and P5 indicated that, “It’s a long story and hard to explain.” This latter response may reflect significant challenges or language-based challenges to sharing their experience. Another possibility is a lack of willingness to share more information due to emotional factors; P5 declined to elaborate. See Table 2 for quality of life results and Supplementary Materials Table 6 for aggregated data.

**Table 2.**
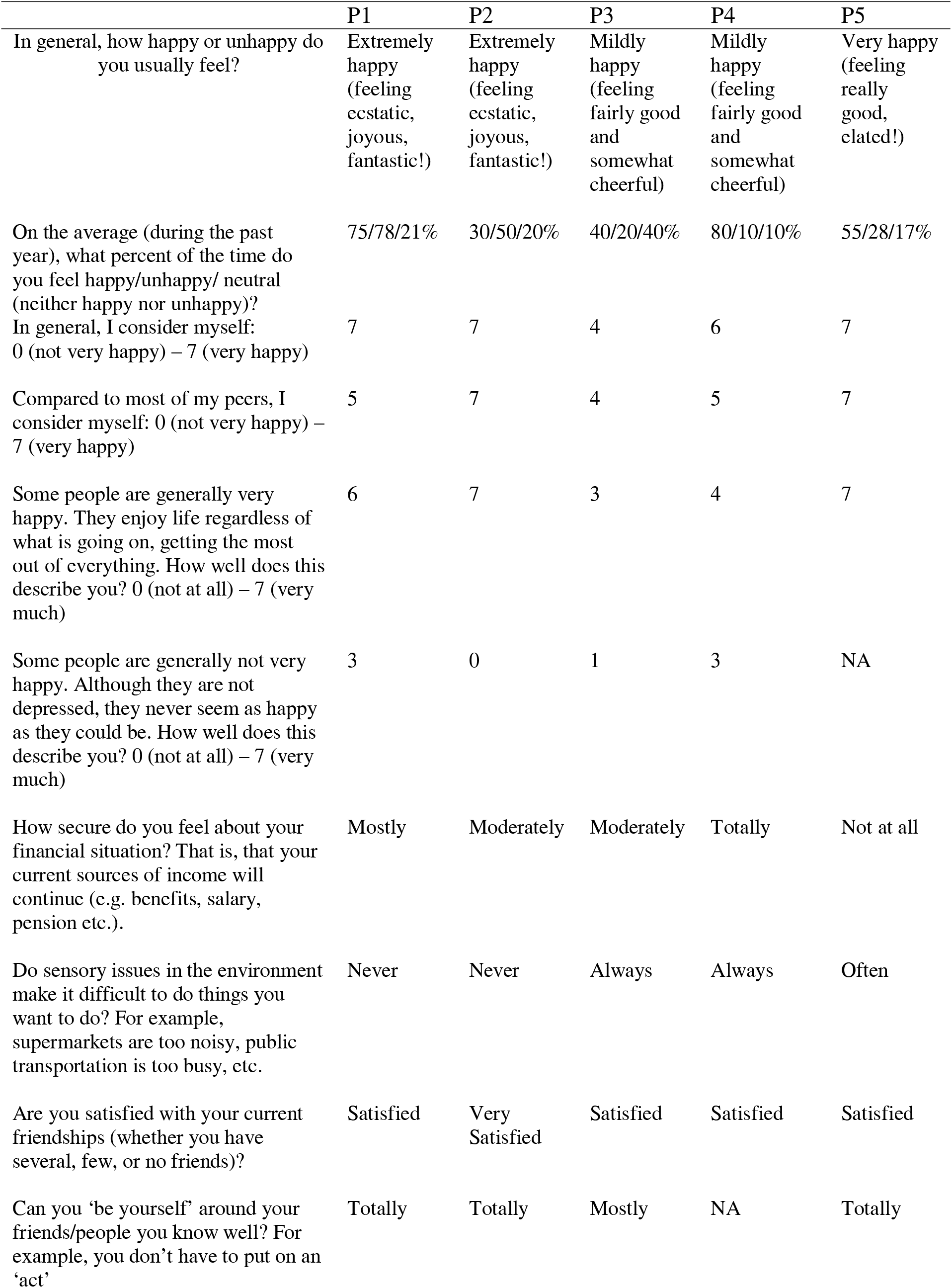
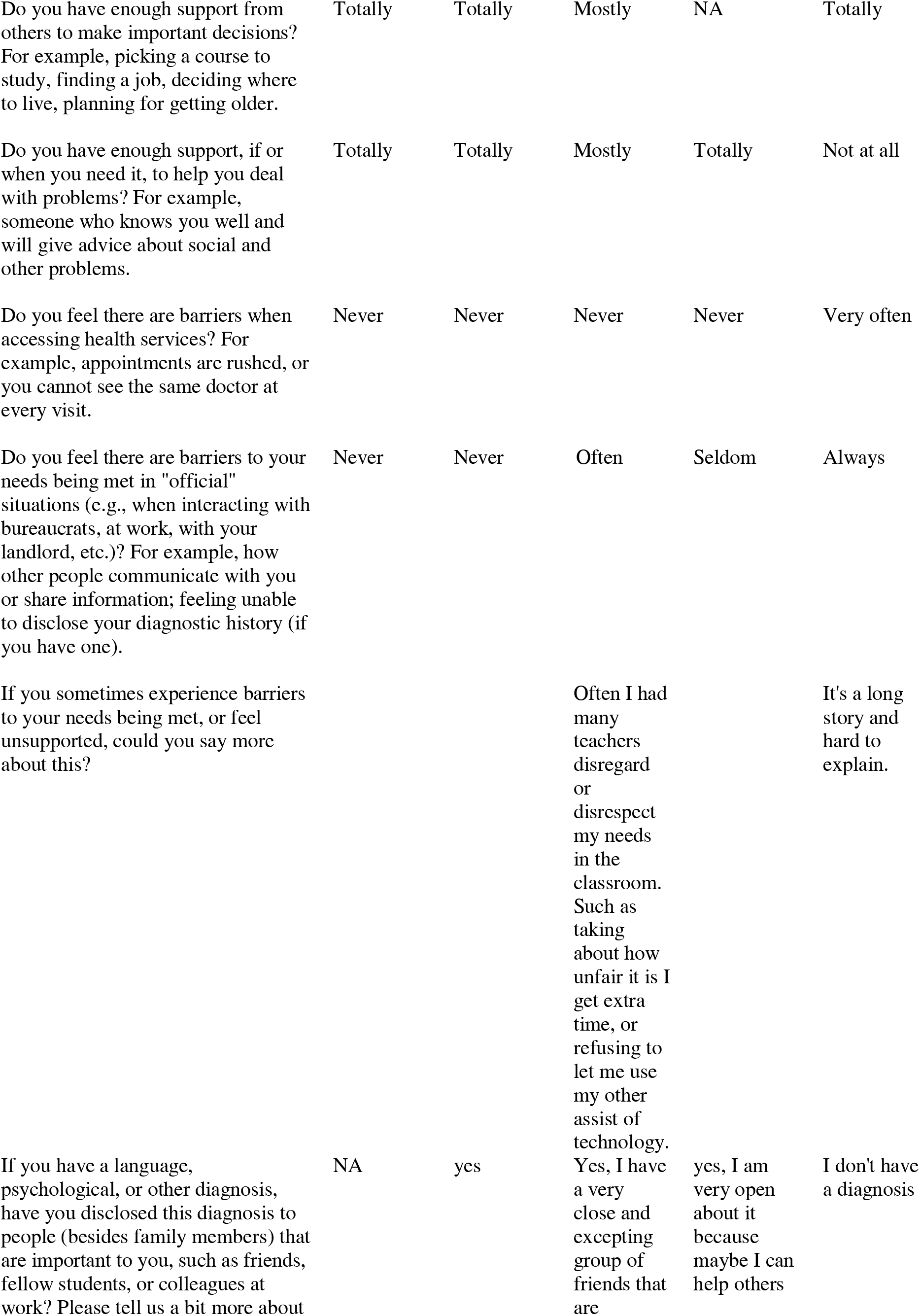
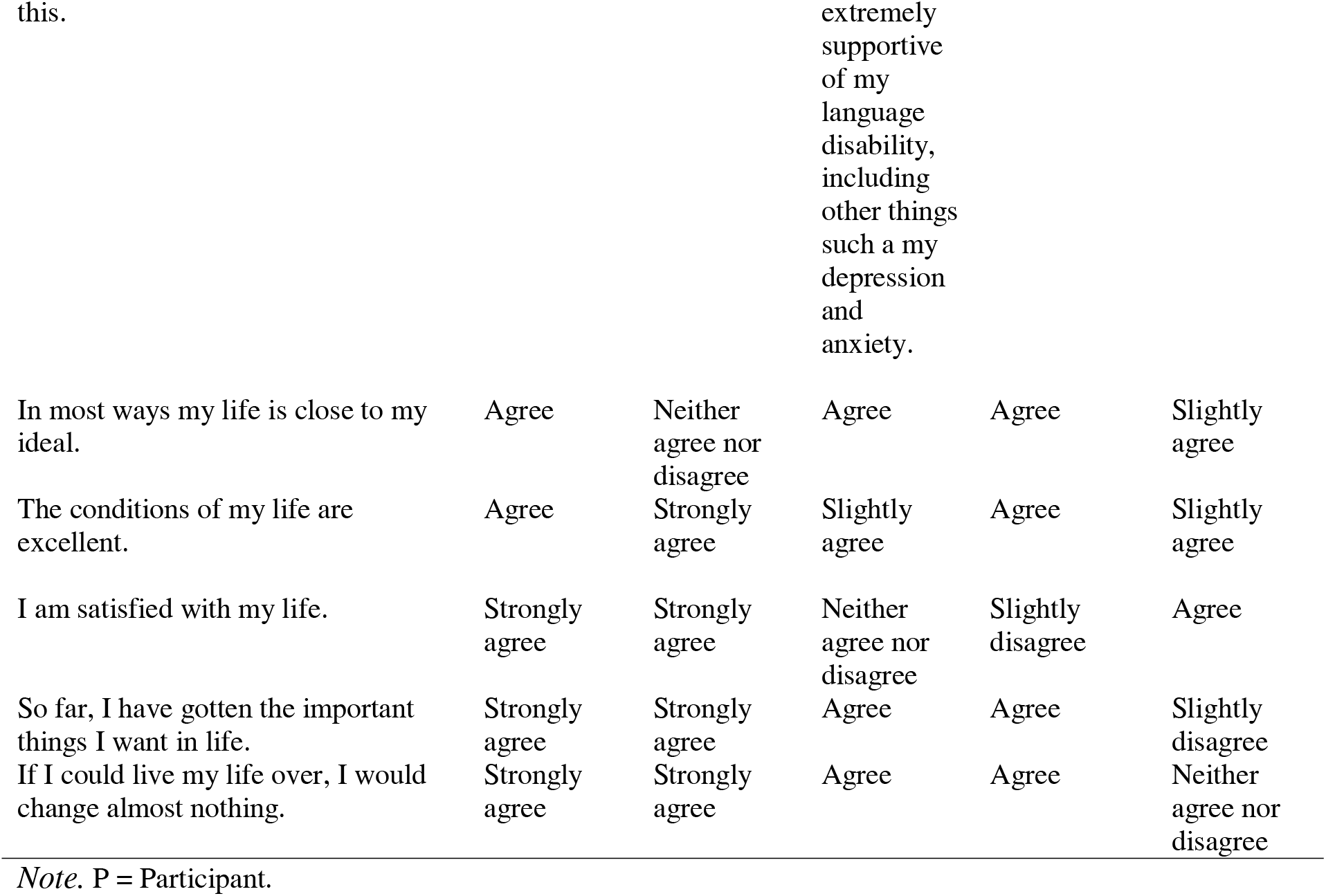
By-participant quality of life and life satisfaction ratings.

### Language and Quality of Life

There were no apparent links between DLD group CELF-5 scores and quality of life measures in the current sample, though it is possible that associations between CELF-5 scores and self-report ratings of happiness, barriers to quality of life, and general life satisfaction may be observed in a larger sample (see CELF-5 results in Supplementary Materials Table 7). There was evidence of several possible links between grammaticality judgement À and quality of life. Relatively higher À scores appeared to be associated with lower ratings of how happy you usually feel, lower ratings of being generally very happy, lower ratings of the ability to ‘be yourself’ around others, and with higher ratings of sensory issues in the environment. Associations between À and being generally very happy and sensory issues in the environment may also be observed in a larger sample. See Table 1 for by-participant language performance, Table 2 for by-participant self-reported quality of life, and Figure 1 for visualization of possible links between À and self-reported quality of life, as well as Supplementary Materials Table 7 for additional results.

**Figure 1.**
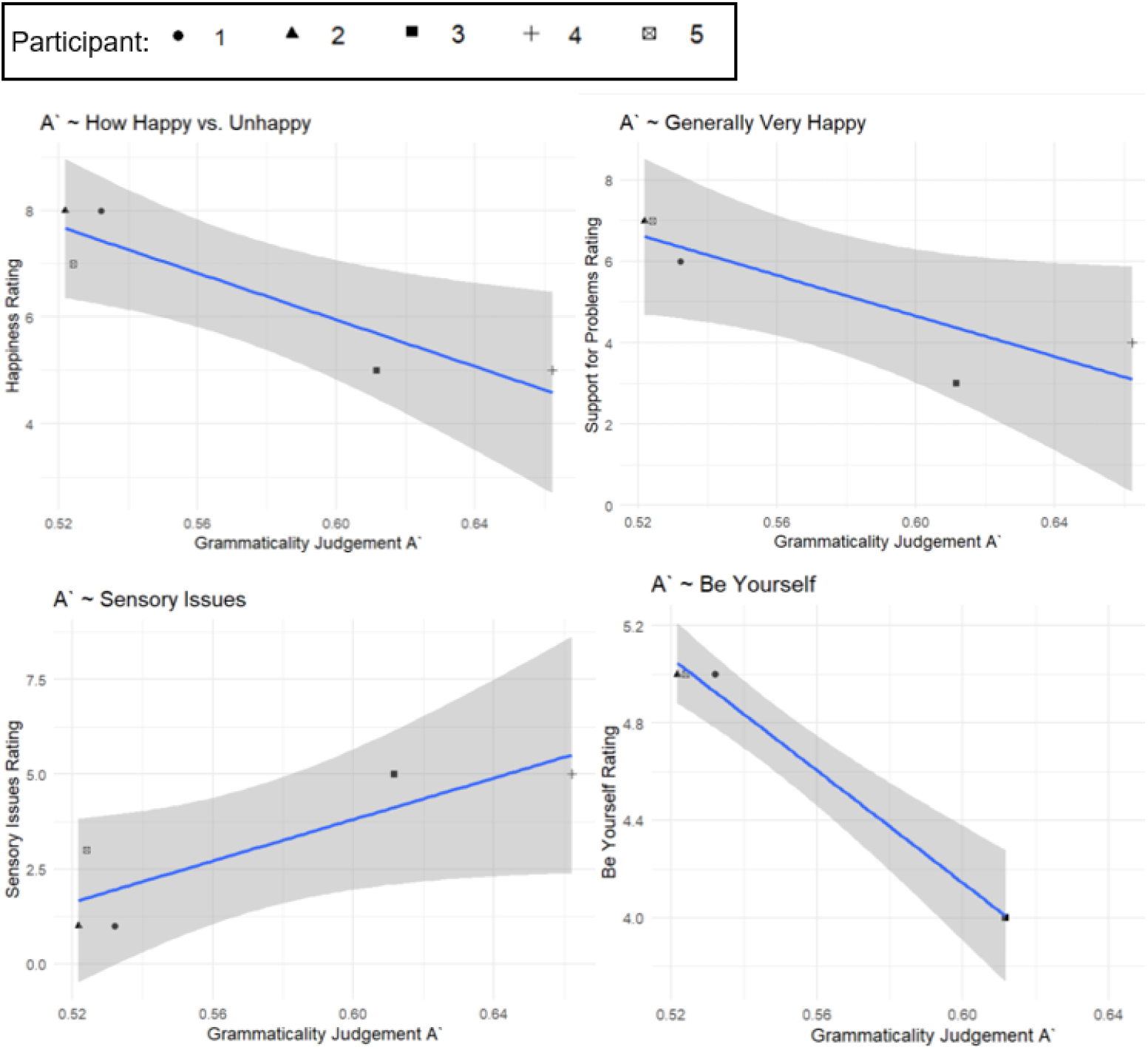
Visualization of possible links between À and self-reported quality of life: how happy versus unhappy you usually feel; generally feeling very happy; sensory issues in the environment making it difficult to function; and ability to ‘be yourself’ for DLD participants.

### Other Risk Factors and Quality of Life

#### Nonverbal ability

Three DLD participants completed the matrix reasoning scale from the WASI-II and all participants had scores within 1.5 standard deviations of the mean (Table 1). P1 had a *t*-score in the average range and P3 had a *t*-score more than one standard deviation above the average range. P1 reported relatively strong quality of life, though somewhat neutral feelings about being generally *not* very happy, and P3 reported lower feelings of happiness, often experiencing barriers in official situations (e.g., accessing accommodations in the classroom), and neutral feelings about general life satisfaction (Table 2). P5 had a *t*-score more than one standard deviation below the mean and reported more barriers and less support on quality of life measures, as well as neutral feelings about life satisfaction (Table 2).

#### Socioeconomic Status

Maternal education and family income were relatively high for all DLD participants (Supplementary Materials Table 2). For P1, maternal highest education and family income were slightly lower than other DLD participants (High school or equivalency versus Associate degree and higher; $75,000-99,000 compared to $100,000 and greater; Supplementary Materials Table 5). This participant reported relatively high quality of life across measures (Table 2).

#### Health

Three DLD participants (P2, P3, and P4) had remarkable health histories. P2 had slightly low birth weight and received brief treatment in the neonatal intensive care unit, P3 had a large head circumference and blood sugar issues, and all three participants experienced Hyperbilirubinemia (jaundice). Participants P3 and P4 had feeding issues and irritability as infants and current ADHD diagnoses, and P3 reportedly had excessive clumsiness (Supplementary Materials Table 3). These participants fell in the middle range on most quality of life measures, and P3 additionally reported barriers related to their educational accommodations (e.g., extra time on tests, assistive technology; Table 2).

## Discussion

This preliminary study explores self-reported quality of life in four adolescents and one young adult with DLD, and the role of language and other risk factors in quality of life. This study addresses recent calls for research on neurodevelopmental conditions across the lifespan and research describing *self-reported* lived experiences of individuals with neurodevelopmental conditions (i.e., neurodiversity-informed approaches), particularly measures that are known to be associated with mental health and wellbeing (Antolini & Colizzi, 2023; Connel at al., 2014; Eigsti et al., 2022; Georgiades & Kasari, 2018; Hobson et al., 2024; e.g., Orrego et al., 2023).

### Self-reported Quality of Life in DLD

Descriptive findings indicated generally good self-reported quality of life in DLD, consistent with self-report findings from Records et al. (1992). In particular, the DLD group generally reported considering themselves a happy person and having strong support from others in making important decisions. Yet, P2, P3, and P5 reported *feeling* happy a relatively low percentage of the time, and P3, P4, and P5 reported experiencing specific barriers in their environments, such as accessing accommodations. These barriers may be associated poorer wellbeing in specific settings, such as those optimized for neurotypical individuals like the regular education classroom (Hobson et al., 2024). For P4, the impacts of sensory barriers in their environment may have been mitigated by their report of disclosing their diagnosis and feeling well supported when dealing with problems, or these barriers may have been limited to non-educational/vocational contexts, such as the grocery store or public transportation. In contrast, P3 and P5 reported less support and experiencing additional barriers, such as barriers in educational contexts. Thus, self-reported quality of life should be examined in context-specific approaches in future research to disentangle barriers in educational versus other settings and the degree to which barriers may be mitigated by support systems.

In contrast to other participants, P5 reported having concerningly little support to deal with problems and experiencing barriers in official situations, including those associated with their higher education experiences (Supplementary Materials Table 3; e.g., accessing health services, interactions at school; see Orrego et al., 2023 for converging evidence). In response to our question probing for further information about these barriers, P5 indicated, “It’s a long story and hard to explain,” (Table 2) which may further suggest language- or communication-based challenges in describing these barriers, though they declined to elaborate. P5 was the only participant in a transitional phase, defined as educational placements and jobs in the first two years after compulsory education (Conti-Ramsden & Durkin, 2012). Individuals with DLD who are in transitional phases have reported feeling *more supported* than their NT peers by their educational system and by their peers in prior work, though individuals with DLD are less likely to be in higher education settings than peers without DLD (∼7% DLD versus ∼78% NT; Conti-Ramsden & Durkin, 2012). Interestingly, P5 reported not having a formal diagnosis, which may have limited their ability to access accommodations needed to feel successful and supported in this transitional phase. Further examining barriers to accommodations noted by P5 and P3 may shed additional light on which individuals with DLD end up in higher education settings and what supports ensure their wellbeing in these settings.

### Language and Quality of Life

There were no apparent links between performance on the standardized language measure, CELF-5, and self-reported quality of life. Yet, it is possible that relationships with happiness, barriers, and general life satisfaction may be identified in a larger, or potentially older, sample (e.g., Conti-Ramsden et al., 2013). Preliminary descriptive evidence suggested possible links between performance on the grammaticality judgement task, which tests this key area of weakness in DLD, and self-reported quality of life. Relatively better performance appeared to reflect lower ratings of happiness and the ability to ‘be yourself’ and with experiencing more sensory-based barriers to functioning in their environment. These patterns suggest that, in our small sample, individuals with DLD who had relatively better language performance reported poorer quality of life on some measures. One possible speculation is that DLD participants with relatively better language skills engage in more verbally mediated self-talk than DLD participants with poorer language skills, and this self-talk, or inner-monologue, involves self-imposed pressure or expectations (e.g., verbal mediation is associated with slower initiation of goal-oriented tasks in DLD; Larson et al., 2019). For instance, the camouflaging literature suggests that masking one’s challenges in social participation is associated with anxiety and depression in neurotypical and autistic individuals (Bargiela et al., 2016; Bernardin et al., 2021; Hull et al., 2017). Masking likely involves a language-mediated inner-monologue that guides such behavior and may underlie the link between language and poorer quality of life in some cases, particularly in environments optimized for neurotypical individuals (Hobson et al., 2024). Though speculative, this interpretation is supported by the apparent link between language and the ability to ‘be yourself’ and should be examined in future research. It is also possible that individuals with DLD who have relatively stronger language skills engage in more neurotypical environments with less support, leading to negative effects on wellbeing. This interpretation is supported by the negative effects of limited supports in high school reported by P3 and in college reported by P5.

The current descriptive evidence converged with other studies indicating a role for language in quality of life, though the directionality of this association should be further examined (Eadie et al., 2018; Haukedal et al., 2023). Eadie et al. (2018) demonstrated that relatively better CELF scores at seven years of age predicted better quality of life at nine years of age, yet this study also demonstrated that mild versus moderate DLD status was not differentially associated with quality of life. Haukedal et al. (2023) reported a positive concurrent association between CELF scores and quality of life in children. In contrast, McGregor et al. (2023) suggested that parent-reported outcomes were associated with risk factors, but not standardized language scores, and Durkin et al. (2012) found an association between CELF scores and academic, but not social-emotional, outcomes in adolescents with DLD or autism and co-occurring language impairment. Notably, most of these studies involve younger participants and *parent-report* rather than *self-report* measures. In a follow-up study of a DLD-only adolescent sample, Conti-Ramsden et al. (2014) reported positive associations between CELF scores and several *self-* and *teacher-* report measures of quality of life, including prosocial behavior and emotional problems. Taken together, there may be nuanced associations between language and quality of life in individuals with DLD which vary depending on *self-* versus *parent-* report, the age or developmental phase of participants, and the particular contexts in question.

### Risk Factors and Quality of Life

Risk factors explored in the current study included nonverbal ability, socioeconomic status, and health history in the DLD group. Descriptive findings supported the possibility that each of these factors contributed to quality of life. First, the participant with the lowest nonverbal ability scores reported more barriers, less support, and more neutral feelings about life satisfaction than other DLD participants. This participant, P5, was in college and receiving accommodations that were reportedly “not very helpful” (Supplementary Materials Table 3). This individual’s experiences are critically important to understand given the paucity of research describing risk factors and quality of life in individuals with DLD who are in transitional phases after high school. Individuals with DLD in transitional phases are more likely to experience a lack of continuity in clinical care and engage in a variety of new experiences that tax their abilities (e.g., living independently, first jobs, college). Somewhat contrastingly, the participant with the highest nonverbal ability scores, P3, reported somewhat low feelings of happiness and life satisfaction, as well as barriers in official situations. Yet, similar to findings for language performance, individuals with DLD who have relatively stronger cognitive skills may be required to function in neurotypical environments with less support, which may be associated with negative effects on wellbeing similar to those observed for P5 (Hobson et al., 2024).

Convergingly, McGregor et al. (2023) found that cumulative risk across multiple factors, including nonverbal abilities, was relevant to functional outcomes in a sample of children with DLD (see also Toseeb et al., 2023). There is also evidence that nonverbal abilities are associated with prosocial behavior (e.g., helping others, social confidence) in adolescents with DLD (Conti-Ramsden et al., 2013). Prosocial behavior may be particularly important to establishing meaningful relationships for individuals with communication deficits. These skills are also essential for effectively self-advocating for supports in contexts that have been structured for neurotypical individuals; an area in which P5 was experiencing barriers. Taken together, nonverbal abilities may be one of multiple factors contributing quality of life that should be further examined.

Second, P1, whose family income and mother’s education were lower than other DLD participants, reported relatively high quality of life. This individual presented with the lowest CELF-5 scores and had a history of receiving multiple special education services, including speech-language services. These descriptive findings are consistent with associations between socioeconomic factors and standardized assessment performance and academic achievement (e.g., Sirin, 2005) and may suggest that socioeconomic factors and associations between socioeconomic factors and academic achievement do not negatively affect quality of life. However, this participant was experiencing few other risk factors identified in prior work, was in high school rather than a later transitional phase, and was receiving multiple supports at school. Thus, future work should test the degree to which socioeconomic status is associated with quality of life *in the absence of* other risk factors (e.g., low nonverbal abilities, poor health; Conti-Ramsden et al., 2013; McGregor et al., 2023; Toseeb et al., 2023).

Finally, the three participants, P2, P3, and P4, with remarkable health histories fell in the middle-range on quality of life measures. This preliminary finding suggests the possibility of a role for health factors in quality of life, albeit an attenuated role in the absence of other risk factors. P3 reported significant barriers to education and had a health history remarkable for Hyperbilirubinemia (jaundice), large head circumference, feeding issues, and irritability as an infant. Although their health history did not appear to be more significant than P2 and P4, their experiences suggest the possibility of educational barriers for individuals with DLD who experience health concerns in infancy. Future work should further examine the role of early health history in quality of life, and how this role varies depending on age and other risk factors.

Collectively, the current descriptive findings align with studies reporting that nonverbal abilities, socioeconomic status, and health history are factors in quality of life for individuals with DLD (e.g., Conti-Ramsden et al., 2013; McGregor et al., 2023; Toseeb et al., 2023), as well as studies indicating quality of life challenges across physical, emotional, and educational domains (Eadie et al., 2018; Haukedal et al., 2023; Le et al., 2021). A preliminary aim for future research derived from the current findings, when considered alongside prior work, is to test how risk factors in particular quality of life challenges may vary depending on age and environmental demands (e.g., Orrego et al., 2023). For instance, the college-age participant, P5, reported greater environmental barriers, less support, and poorer general life satisfaction than other DLD participants. It would be useful to understand how educational barriers P3 reported experiencing in high school and potential associations between these barriers and quality of life unfold over time. Clearly, the clinical course and lived experiences of individuals DLD in and beyond high school should be characterized to understand factors contributing to wellbeing and to approach these questions from a neurodiversity-informed perspective.

### Limitations

Given the scope of this preliminary study, findings should be interpreted with caution and as exploratory directions for future research. A primary limitation is the small sample size and descriptive nature of this work. Additionally, while self-report measures were rich in information, there are known limitations with self-report measures, such as the possibility that DLD participants had challenges understanding the questions. However, no participants reported challenges with completing any measures in the current study.

### Implications and Future Directions

This preliminary study contributes exploratory evidence on *self-reported* quality of life and factors associated with quality of life in four adolescents and one adult with DLD. Overall quality of life did not appear to be remarkably poor in DLD. Yet, the descriptive evidence suggests barriers in educational and official contexts, such as receiving appropriate accommodations in school and in accessing health services, particularly for our adult participant who was in college. Language performance on a task that measured an area of disproportionate weakness in DLD, morphosyntax, had possible links with quality of life measures, including happiness, the ability to ‘be yourself,’ and experiencing sensory-based barriers to functioning in your environment. These associations suggested that individuals with DLD and relatively better language performance reported poorer quality of life. The current preliminary study suggests a larger role for nonverbal abilities than socioeconomic status and health history as risk factors in quality of life, as well as the possibility that cumulative risk and life phase have a larger impact on quality of life than individual risk factors. Taken together, this exploratory evidence provides direction for future, neurodiversity-informed research to examine *self-reported* quality of life and how it varies depending on language skills and risk factors. These findings may also represent considerations for clinicians to explore when working with individuals with DLD to identify functional challenges that may impact mental health and wellbeing.

## Data Availability

All data produced in the present study are available upon reasonable request to the authors

## Acknowledgements

I gratefully acknowledge funding from the University of Connecticut Institute for the Brain and Cognitive Sciences (Seed Grant, PI Larson) and the National Institutes of Health R01MH112687-01A1 (PIs Eigsti and Fein) and would like to thank the research assistants and study participants and their families who made this research possible, as well as Elizabeth Kelley, Laura Morett, and Robin Karlin for their helpful feedback.

## Supplemental Materials

This file contains additional descriptive and aggregated data, additional methods on descriptive approach to analysis, and comparisons between language performance and quality of life.

